# A comparison of prevalence estimates of smoking, alternative nicotine and alcohol use in Great Britain collected via telephone versus face-to-face: Smoking and Alcohol Toolkit surveys

**DOI:** 10.1101/2024.07.30.24311204

**Authors:** Vera Helen Buss, Loren Kock, Harry Tattan-Birch, Sarah E. Jackson, Lion Shahab, Jamie Brown

**Affiliations:** Department of Behavioural Science and Health, University College London, UK; SPECTRUM Research Consortium; Department of Psychiatry, University of Vermont, US

**Keywords:** Cross-Sectional Studies, Survey Methodology, Demographics, Tobacco Use, Nicotine, Alcohol Drinking

## Abstract

**Background and Aims:** Due to the COVID-19 pandemic, the survey mode of the Smoking and Alcohol Toolkit Study, a long-running repeat cross-sectional survey, had to change from face-to-face to telephone interviews. This study aimed to assess similarities and differences in sociodemographic, smoking, alternative nicotine and alcohol use estimates between the two survey modes, to understand the potential impacts of this change in methodology on prevalence estimates and trends over time.

**Design:** After COVID-19 restrictions were lifted, we conducted parallel telephone and face-to-face household surveys in March 2022 and in January to March 2024, using a hybrid of random and quota sampling. Data from both years were aggregated.

**Setting and Participants:** People aged 16+ years living in private households in Great Britain.

**Measurements:** Sociodemographic characteristics, nicotine and alcohol use related estimates and their 95% CIs — unweighted and weighted — collected via telephone versus face-to-face in a household.

**Findings:** In the unweighted analyses, the telephone sample included slightly younger and less socioeconomically advantaged groups than the face-to-face sample. After the samples were weighted, estimates of sociodemographic characteristics and nicotine and alcohol use were generally consistent across methodologies, including daily cigarette smoking (face-to-face: 11.1% [10.1-12.1] vs. telephone: 10.6% [9.5-11.7]), non-daily cigarette smoking (face-to-face: 2.7% [2.2-3.3] vs. telephone: 3.4% [2.8-4.1]), and e-cigarette use among people who smoke (face-to-face: 27.0% [23.5-30.5] vs. telephone: 29.3% [25.4-33.3]). However — compared with telephone participants — a lower proportion of face-to-face participants reported currently using e-cigarettes (face-to-face: 6.4% [5.6-7.1] vs. telephone: 10.4% [9.3-11.5]), and a higher proportion reported never drinking alcohol (face-to-face: 31.1% [29.7-32.5] vs. telephone: 25.0% [23.5-26.5]) and never having 6 or more standard drinks on one occasion (face-to-face: 46.6% [44.7-48.5] vs. telephone: 40.2% [38.4-42.1]). More participants provided “don’t know” or “refused” responses in the telephone compared with the face-to-face interview, including in response to questions about tobacco use, e-cigarette device type, and the number of standard drinks on a typical day.

**Conclusions:** Face-to-face and telephone surveys generally yield similar estimates of nicotine and alcohol use. However, there may be some underreporting of vaping and drinking in a face-to-face survey conducted in the home compared with telephone.

## Introduction

The Smoking and Alcohol Toolkit Study (STS/ATS) is a long-running monthly representative household survey. It started in England in November 2006 and expanded to cover all of Great Britain (i.e., England, Wales and Scotland) from October 2020 (1-3). Up to February 2020, the survey was conducted face-to-face, but due to the COVID-19 pandemic, it changed to telephone surveys from April 2020. This mode change could have impacted estimates generated through the survey, which is a particular issue when comparing trends over time. The current study aimed to compare key indicators of nicotine and alcohol use collected via telephone versus face-to-face interviews.

A previous study comparing both data collection modes conducted in March 2022 as part of the STS/ATS showed that more young people participated in the face-to-face compared to the telephone survey, but otherwise sociodemographic characteristics were similar (4). After data were weighted, differences were found in estimates for never smoking and having quit more than a year ago, but when those two were combined, the estimates were similar (4). For alcohol use, there were differences in those reporting not drinking, with a higher estimate found in the face-to-face than the telephone survey (4). Overall, most estimates were comparable between the two data collection modes (4).

We conducted another parallel wave of face-to-face and telephone data collection in STS/ATS in the first quarter of 2024. The current study aimed to update the previous analysis, combining the March 2022 data with the new data collected in the first quarter of 2024. In addition to the variables assessed in the previous study, the present study also included estimates of the use of various nicotine products in the general population, as the use of novel nicotine products has increased among people who do not smoke in recent years (5).

## Methods

### Sample and recruitment

Data were drawn from the STS/ATS, a monthly repeat cross-sectional study of a representative sample of approximately 2,450 people aged 16 years or over living in Great Britain (3). The sampling method consists of a hybrid of random location and quota sampling (1). Since April 2020, the interviews are routinely conducted via computer-assisted telephone interviewing. In March 2022 and February 2024, 2,607 and 2,375 individuals were interviewed respectively. For this study, the same survey questions were asked to a sample of 2,064 and 2,317 individuals, respectively, via computer-assisted personal interviewing (i.e., face-to-face) in March 2022 and between January and March 2024 (21% in January, 65% in February, and 14% in March). The University College London Ethics Committee granted ethical approval for the STS/ATS (ID 0498/001).

### Measures

This study included measures on sociodemographic characteristics (age, gender, and social grade (6)), smoking status, quitting-related measures for those who smoked in the past year, other nicotine use among those who smoked in the past year and across the total population, e-cigarette device used, and alcohol consumption (questions of the Alcohol Use Disorders Identification Test – Consumption [AUDIT-C]) (7), and whether people scored ≥5 on the AUDIT-C, which is operationalised as increasing and higher risk drinking (8)). More details on how the measures were derived are provided in the supplementary material.

### Analysis

For both survey modes, we combined the data collected in 2022 and 2024. For the 2024 telephone sample, we used data collected in February 2024 (when the majority of the face-to-face data were collected). First, we reported the number of missing values for relevant variables for both survey modes. Second, we compared unweighted and weighted estimates and their 95% confidence intervals (CIs) between both survey modes for sociodemographic characteristics, smoking, alternative nicotine and alcohol use related measures. Data weighting was performed using raking (9), to adjust estimates to accurately represent the population of Great Britain in terms of sociodemographic characteristics, including age, gender, social grade, working status, prevalence of children in the household, and region (1).

There were several sensitivity analyses. First, we compared the two datasets from the different modes collected in 2024 only. Second, we repeated the weighted analysis using telephone data collected between January and March 2024, given the longer face-to-face data collection period in 2024. For this analysis, participants’ weights were rescaled according to the percentage of face-to-face data collected in each respective month (see supplementary file). Third, we investigated differences between modes for e-cigarette use in more detail by stratifying e-cigarette use by age. Forth, we calculated the number and proportion of participants who reported e-cigarette use in response to each of the different survey questions that contributed to the derived variable “e-cigarette use in total population”.

## Results

The face-to-face sample comprised of 4,381 participants and the telephone sample of 4,982 participants. Table 1 shows missing values by variables for both survey modes. These are values that were missing rather than answer responses coded as “don’t know” or “refused”. However, for the variable “AUDIT-C ≥5”, missing values are a combination of participants answering “don’t know” or “refused” to at least one of the AUDIT items, resulting in missing values for the composite score. There were no missing values for the remaining variables due to the way they were derived.

**Table 1:** Missing values during face-to-face (F2F) and telephone (Tel) data collection.

| Variable | F2F missing values |  | Tel missing values |  |
| --- | --- | --- | --- | --- |
|  | n | % (95% CI) | n | % (95% CI) |
| Age | 0 | 0 (0-0) | 0 | 0 (0-0) |
| Gender | 8 | 0.2 (0.1-0.3) | 24 | 0.5 (0.3-0.7) |
| Social grade | 0 | 0 (0-0) | 0 | 0 (0-0) |
| Smoking status | 0 | 0 (0-0) | 0 | 0 (0-0) |
| AUDIT 1 | 0 | 0 (0-0) | 0 | 0 (0-0) |
| AUDIT 2 and 3 (N<sub>F2F</sub>=2973, N<sub>Tel</sub>=3774)<sup>1</sup> | 23 | 0.8 (0.5-1.1) | 0 | 0 (0-0) |
| AUDIT-C ≥5 | 49 | 1.1 (0.8-1.4) | 220 | 4.4 (3.8-5.0) |
| Quit attempt in past year (N<sub>F2F</sub>=702, N<sub>Tel</sub>=794)<sup>2</sup> | 52 | 7.4 (5.5-9.3) | 41 | 5.2 (3.7-6.7) |
| E-cigarette device used (N<sub>F2F</sub>=259, N<sub>Tel</sub>=417)<sup>3</sup> | 21 | 8.1 (4.8-11.4) | 19 | 4.6 (2.6-6.6) |

<sup>1</sup> only assessed for people who reported drinking alcohol for AUDIT 1 question.
<sup>2</sup> only assessed for people who reported smoking in the past year.
<sup>3</sup> only assessed for people who reported using e-cigarettes.

### Sociodemographic characteristics

In the unweighted data, face-to-face survey participants were slightly younger than telephone survey participants (Figure 1). There was also a slight difference according to social grade, with fewer people in social grades AB and C1 and fewer in C2 and D in the face-to-face compared to the telephone survey. After weighting, both samples matched the population targets for England by design (supplementary Table S1).

**Figure 1:**
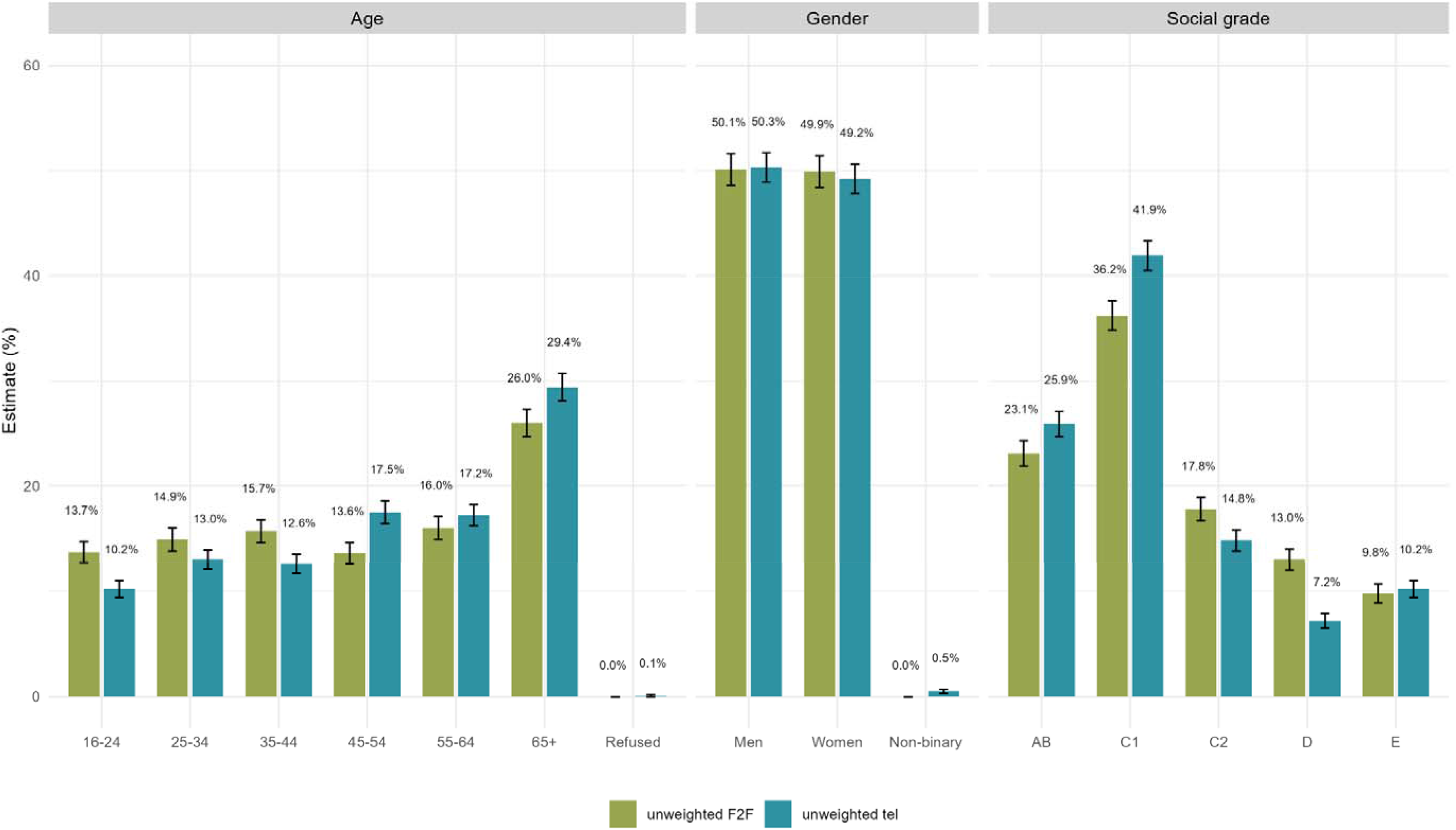
Unweighted sociodemographic estimates from face-to-face (F2F) and telephone (Tel) data collection (N_F2F_=4381 and N_Tel_=4982). Error bars represent 95% confidence intervals.

### Nicotine use

After applying the sampling weights, estimates for most nicotine use variables were comparable across survey modes (Figure 2 and supplementary Table S2). Among the differences were that the face-to-face sample reported lower proportions of having stopped smoking over a year ago and higher proportions of never smoking relative to the telephone sample. Less people responded “don’t know” in the face-to-face than the telephone interviews regarding their tobacco use.

**Figure 2:**
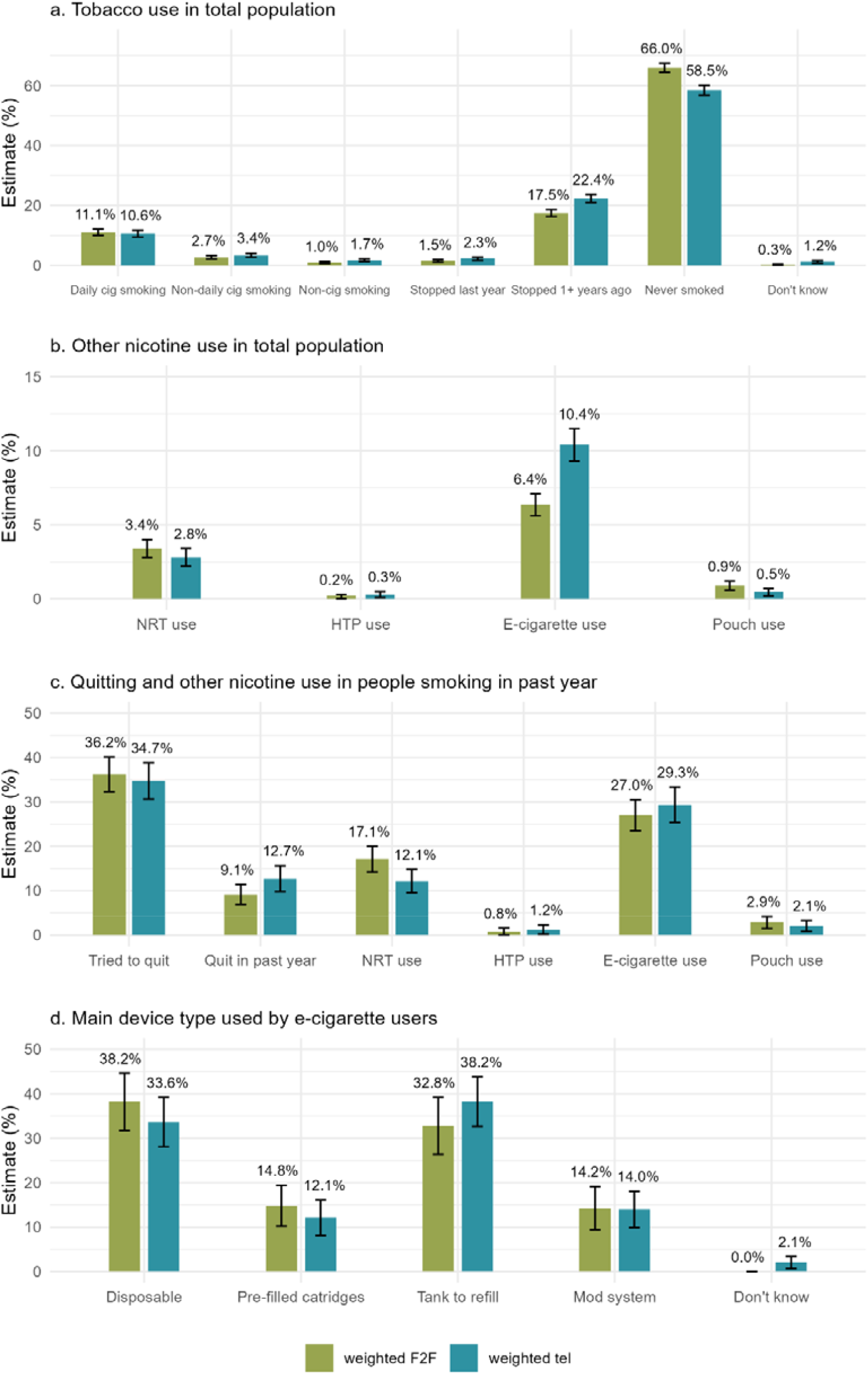
Weighted estimates for smoking and use of other nicotine products from face-to-face (F2F) and telephone (Tel) data collection (N_F2F_=4381 and N_Tel_=4982; people smoking in past year: N_F2F_=702 and N_Tel_=794; e-cigarette users: N_F2F_=259 and N_Tel_=417). Error bars represent 95% confidence intervals. Abbreviations: cig, cigarette; NRT, nicotine replacement therapy; HTP, heated tobacco product; pre-filled cartridges, rechargeable device with pre-filled cartridges; tank to refill, rechargeable device with tank to refill.

A lower proportion of the face-to-face sample than the telephone sample reported using e-cigarettes. During the telephone interviews, some e-cigarette users reported not knowing what device they usually used, while no one provided this response during the face-to-face interviews.

### Alcohol use

Similar to nicotine use, weighted estimates of most alcohol use variables were comparable across survey modes (Figure 3 and supplementary Table S3). A higher proportion of those surveyed via face-to-face compared to telephone reported never drinking at all (AUDIT 1) and never having 6 or more standard drinks on one occasion (AUDIT 3). This resulted in a lower prevalence of increasing and higher risk drinking (AUDIT-C ≥5) among the face-to-face sample (face-to-face: 25.3%, 95% CI: 23.9-26.3 vs. telephone: 30.4%, 95% CI: 28.9-32.0). Compared to the face-to-face sample, a higher proportion of the telephone sample provided “don’t know” or “refused” responses.

**Figure 3:**
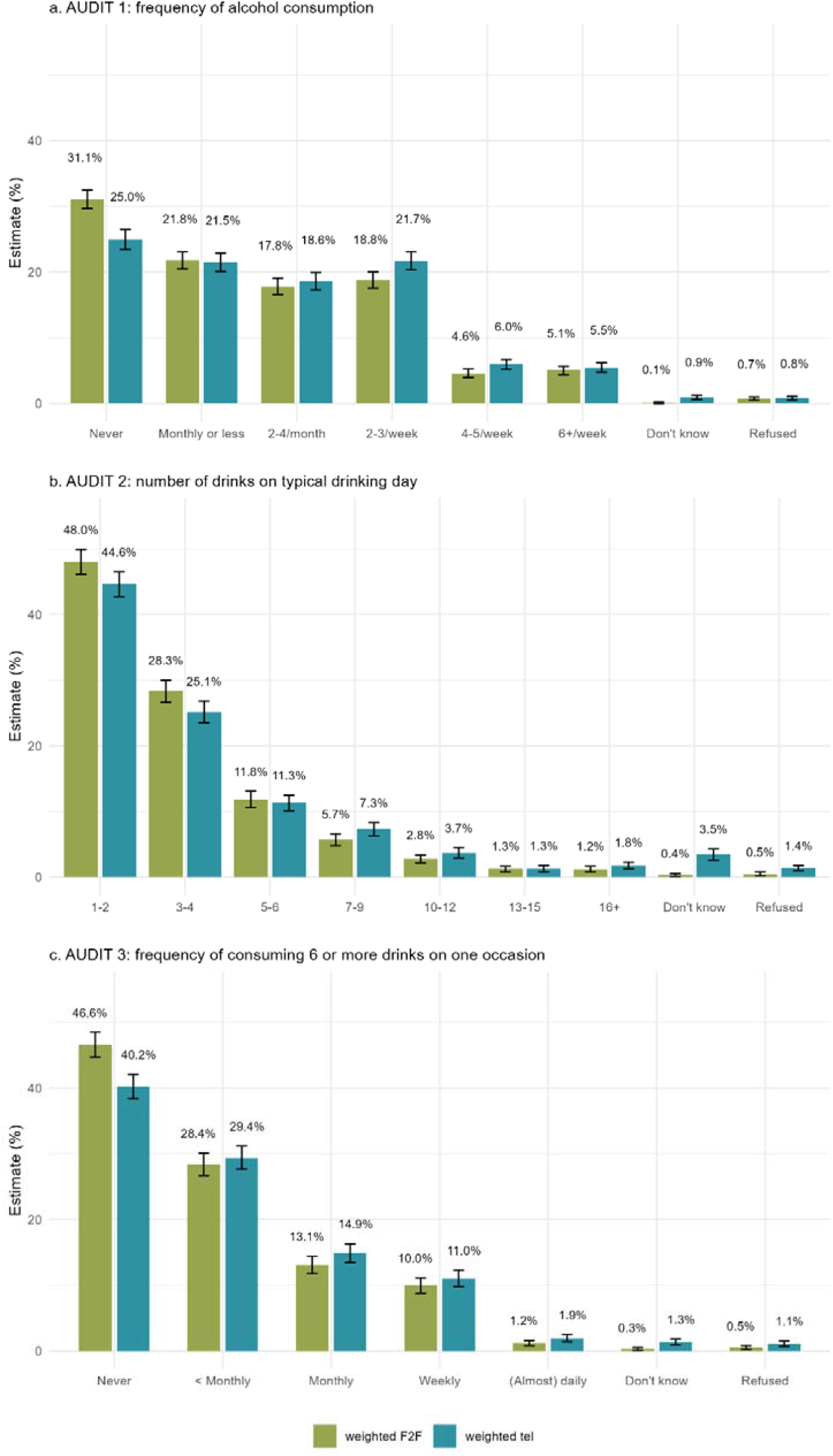
Estimates for alcohol use from face-to-face (F2F) and telephone (Tel) data collection (N_F2F_=4381 and N_Tel_=4982). Error bars represent 95% confidence intervals.

### Sensitivity analyses

Results were not materially altered when using telephone data from January to March 2024 data to match face-to-face distribution of data collection instead (supplementary Tables S8-S10). In the age-stratified analysis for e-cigarette use (supplementary Table S11), the age groups 16-24 years, 25-34 years, and 35-44 years had higher e-cigarette use estimates in the weighted and unweighted analyses in the telephone compared with the face-to-face survey, while the values for the other age groups were comparable across the two survey modes. By comparing the number and proportion of positive responses to each variable that contributed to the derived variable “current e-cigarette use in the total population” (supplementary Table S12), the largest difference between survey modes was for the question “Can I check, are you using any of the following?” (unweighted: n_F2F_=110 of 3667, 3.0% [95% CI: 2.4-3.6] vs. n_Tel_=209 of 4131, 5.1% [95% CI: 4.4-5.8]), which was asked to those who did not smoke in the past year while the other variables were asked to those who currently smoked or smoked in the past year.

## Discussion

Overall, most estimates were comparable across the two survey modes. The face-to-face sample included slightly younger and less socioeconomically advantaged people than the telephone sample, which was balanced out through weighting. After weighting, there were only differences in the estimates of the prevalence of never smoking, having stopped smoking over a year ago, overall prevalence of e-cigarette use, never drinking alcohol, and never having 6 or more standard drinks on one occasion. For some variables, there were more “don’t know” or “refused” responses among the telephone compared to the face-to-face sample, including the questions about tobacco use, e-cigarette device type, and the number of standard drinks on a typical day.

These findings are consistent with other research comparing data quality between both survey modes. Generally, differences in mode can affect sample representativeness as well as observed response patterns (10). The first point is less of a concern when data weighting is used as it is routinely applied in the STS/ATS. However, it is possible that people who are not included in our surveys because they are either not reached or refused to participate are systematically different from those who participate (10, 11). The higher “don’t know” response rate in the telephone survey may be due to people putting less effort into answering the questions compared with people in face-to-face surveys (‘satisficing’) (10). Other research has also shown that there is a general tendency towards more “don’t know” responses during telephone compared to face-to-face interviews (12, 13). In face-to-face interviews, the interviewers can use non-verbal communication, potentially increasing engagement and encouraging participants to provide an answer (rather than saying “don’t know”) (10, 12).

We generally observe large month-to-month variations between the rates of never smoking and having quit more than a year ago, even when data are collected using the same survey mode. The reason for the variation could be that people who quit a long time ago and may never have smoked heavily classify themselves as never-smokers on some days and ex-smokers on others. While the prevalence of never smoking was higher in the face-to-face compared to the telephone survey (66.0% vs. 58.5%), the opposite was the case for having stopped over a year ago (17.5% vs. 22.4%). In our analyses, we often combine the two categories to “not smoked in the past year”, in which case these differences in responding would not have a material impact. The differences in alcohol consumption estimates may also be partly due to random variability in responses between never and rare alcohol consumption and partly due to social desirability bias, as the same tendencies were apparent in the 2022 (4) and 2024 datasets.

The present study also found a difference (between 2 to 6 percentage points) in e-cigarette use prevalence in the total population between face-to-face (6.4%) and telephone (10.4%) interviews, which may be driven by differential responses from younger age groups. Other studies also reported some variation in this outcome between survey settings. In the 2021 US National Youth Tobacco Survey, students could complete the survey online during class either in school or at home (14). The proportion of students reporting past-30-day e-cigarette use was about twice as high among those taking the survey at school (15.0%) as among those taking it at home (8.2%) (14). This effect persisted after adjusting for various covariates. The authors attributed at least part of it to response bias — depending on whether the students were among their peers or parents/guardians (14). In a 2019 systematic review, Levy et al. (15) found that online surveys tended to measure higher e-cigarette use rates than telephone or in-person surveys. Since our study showed that the difference in estimates was larger among younger and middle-aged adults, we hypothesise that they may have felt more reluctant to admit to e-cigarettes use in the face-to-face household interviews. The same issue may have led to some underreporting of alcohol consumption in the face-to-face samples. In future, we aim to record whether other people are present in the household while the survey is being completed. Assuming that our results indicate a lower risk of response bias during telephone interviews, our new study methodology may yield more accurate responses.

It is worth noting some limitations of the study. First, the sample sizes may have not been sufficient to detect important differences of some variables. Second, this is the second time we have compared a relatively large number of indicators; therefore, it is possible that some of the highlighted differences may reflect random variation. Third, data were not collected at exactly the same time. We tried to adjust for this by including a sensitivity analysis with telephone data collected between January and March 2024, which is the same timeframe in which the face-to-face survey was collected. The results of this analysis were comparable to that using only February 2024 data.

In summary, face-to-face and telephone surveys generally yielded similar estimates of nicotine and alcohol use. However, there may be some underreporting of vaping and drinking in a face-to-face survey conducted in the home compared with telephone.

## Data Availability

Data are available upon reasonable request.

## Data sharing

Data are available upon reasonable request.

## Acknowledgements

This work was supported by Cancer Research UK (PRCRPG-Nov21\100002) and the UK Prevention Research Partnership (MR/S037519/1), which is funded by the British Heart Foundation, Cancer Research UK, Chief Scientist Office of the Scottish Government Health and Social Care Directorates, Engineering and Physical Sciences Research Council, Economic and Social Research Council, Health and Social Care Research and Development Division (Welsh Government), Medical Research Council, National Institute for Health Research, Natural Environment Research Council, Public Health Agency (Northern Ireland), The Health Foundation and Wellcome.

The study sponsors did not have any role in the collection, analysis, and interpretation of data; in the writing of the report; and in the decision to submit the paper for publication.

## Notes

### Competing Interest Statement

JB has received unrestricted research funding from Pfizer and J&J, who manufacture smoking cessation medications. LS has received honoraria for talks, unrestricted research grants and travel expenses to attend meetings and workshops from manufactures of smoking cessation medications (Pfizer; J&J) and has acted as paid reviewer for grant awarding bodies and as a paid consultant for health care companies. All authors declare no financial links with the tobacco, e-cigarette, or alcohol industry or their representatives.

### Author Declarations

The University College London Ethics Committee granted ethical approval for the Smoking and Alcohol Toolkit Study (ID 0498/001).

