## Supplementary File for "A comparison of prevalence estimates of smoking, alternative nicotine and alcohol use in Great Britain collected via telephone versus face-to-face: Smoking and Alcohol Toolkit surveys"

### 1. Measures

**Age**: categories into 16-24/25-34/35-44/45-54/55-64/65+ years [asked to all]

**Gender:** categorised into men, women, or non-binary [asked to all]

**Social grade**: as a measure of socioeconomic position, categorised into AB (high or intermediate managerial, administrative or professional), C1 (supervisory, clerical and junior managerial, administrative or professional), C2 (skilled manual workers), D (semi and unskilled manual workers), E (state pensioners, casual or lowest grade workers, unemployed with state benefits only).[1] [asked to all]

**Smoking status:** assessed with the following question, which was asked to all,

*"Which of the following best applies to you? Please note we are referring to cigarettes and other kinds of tobacco that you set light to and NOT electronic or 'heat-not-burn' cigarettes.*

1. *I smoke cigarettes (including hand-rolled) every day*
2. *I smoke cigarettes (including hand-rolled), but not every day*
3. *I do not smoke cigarettes at all, but I do smoke tobacco of some kind (eg. Pipe, cigar or shisha)*
4. *I have stopped smoking completely in the last year*
5. *I stopped smoking completely more than a year ago*
6. *I have never been a smoker (i.e. smoked for a year or more)*
7. *Don't know"*

**Smoking in the past year:** binary variable – yes, if one of answer options 1-4 selected above [asked to all]

**Tried to stop smoking in past year:** binary variable – assessed with the following question which was asked to those who smoked in the past year (see above),
*"How many serious attempts to stop smoking have you made in the last 12 months? By serious attempt I mean you decided that you would try to make sure you never smoked again. Please include any attempt that you are currently making and please include any successful attempt made within the last year."*

[1] Collis, D. (2009). Social grade: A classification tool – Bite sized through piece (Ipsos MediaCT, Ipsos Mori). https://www.ipsos.com/en-uk/social-grade

Categorised as tried to stop smoking in the past year if answered with at least one attempt.

**Quit smoking in past year among those who smoked in past year:** binary variable – assessed with the following question, which was asked to all,

*"Which of the following best applies to you? Please note we are referring to cigarettes and other kinds of tobacco that you set light to and NOT electronic or 'heat-not-burn' cigarettes.",*

and the answer response, *"I have stopped smoking completely in the last year"*.

Proportion calculated by diving the number of people with that response by the number of people who smoked in the past year (see above).

**NRT/HTP/e-cigarette/pouch use (in people smoking in the past year or in total population):** assessed with the following questions,

- *"Which, if any, of the following are you currently using to help you cut down the amount you smoke?"* [asked to people who currently smoke]
- *"Do you regularly use any of the following in situations when you are not allowed to smoke?"* [asked to people who currently smoke]
- *"Can I check, are you using any of the following?"* [asked to people who have not smoke in past year]
- *"Can I check, are you using any of the following either to help you stop smoking, to help you cut down or for any other reason at all?"* [asked to people who smoked in past year]

Categorised as using nicotine replacement therapy (NRT) if answered *"Nicotine gum", "Nicotine replacement lozenges\tablets", "Nicotine replacement inhaler", "Nicotine replacement nasal spray", "Nicotine mouthspray"* or *"Nicotine patch"* to any of these questions.

Categorised as using heated tobacco products (HTP) if answered *"Heat-not-burn cigarette (e.g. iQOS with HEETS, heatsticks)"* to any of these questions.

Categorised as using e-cigarettes if answered *"Electronic cigarette or vaping device"* or *"Juul"* to any of these questions.

Categorised as using nicotine pouches if answered *"Tobacco-free nicotine pouch/pod or 'white pouches' that you place on your gum (e.g., Zyn, On!, Nordic Spirit, Velo, Lyft, Skruf)"* to any of these questions.

**E-cigarette device used:** assessed with the following question, which was asked to those who reported using e-cigarettes (see above),

*"Which of the following do you mainly use…?*

1. *A disposable e-cigarette or vaping device (non –rechargeable)*
2. *An e-cigarette or vaping device that uses replaceable pre-filled cartridges (rechargeable)*
3. *An e-cigarette or vaping device with a tank that you refill with liquids (rechargeable)*
4. *A modular system that you refill with liquids (you use your own combination of separate devices: batteries, atomizers, etc.)*
5. *Don't know"*

**AUDIT-C questions**

*"The next few questions form part of a study about consumption of alcohol. We understand that this is a highly sensitive topic and would therefore like to remind you that any information you give me is strictly confidential and will be used for research purposes only. Some questions asked may not necessarily apply to you.*

*These first few questions ask about the alcohol you have drunk in the last 6 months, including about how many standard drinks you have consumed. Please note that 1 standard drink equals 1 unit of alcohol. So, for example, a small glass of wine or a single measure of spirits is 1 standard drink, while a pint of regular beer or lager is equal to 2 standard drinks or 2 units, and a bottle of wine is equal to 9 units. If you are unsure, please ask me to help you work it out.*

*Please be aware that all your answers will be handled confidentially.*

*AUDIT 1. How often do you have a drink containing alcohol?*

i. *Never*
ii. *Monthly or less*
iii. *2 to 4 times a month*
iv. *2 to 3 times a week*
v. *4 to 5 times a week*
vi. *6 or more times a week*
vii. *Don't know*
viii. *Refused*

*AUDIT 2. How many standard drinks containing alcohol do you have on a typical day when you are drinking?*

i. *1 to 2*
ii. *3 to 4*
iii. *5 to 6*
iv. *7 to 9*
v. *10 to 12*
vi. *13 to 15*
vii. *16 or more*
viii. *Don't know*
ix. *Refused*

*AUDIT 3. How often do you have six or more standard drinks on one occasion?*

i. *Never*
ii. *Less than monthly*
iii. *Monthly*
iv. *Weekly*
v. *Daily or almost daily*
vi. *Don't know*
vii. *Refused"*

AUDIT 1 asked to all, AUDIT 2 and AUDIT 3 to those who responded with one of answer options ii to vi to AUDIT 1 (i.e., those who reported alcohol consumption).

**Increasing or higher risk drinking:** asked to all and operationalised as AUDIT-C score of 5 or above

AUDIT 1 – answer options and their respective value for the AUDIT-C score:

- Never: 0
- Monthly or less: 1
- 2 to 4 times a month: 2
- 2 to 3 times a week: 3
- 4 to 5 times a week or 6 or more times a week: 4

AUDIT 2 – answer options and their respective value for the AUDIT-C score:

- 1 to 2: 0
- 3 to 4: 1
- 5 to 6: 2
- 7 to 9: 3
- 10 to 12 or 13 to15: 4

AUDIT 3 – answer options and their respective value for the AUDIT-C score:

- Never: 0
- Less than monthly: 1
- Monthly: 2
- Weekly: 3
- Daily or almost daily: 4

### 2. Unweighted and weighted data (2022 and 2024)

This section includes face-to-face data collected in March 2022 and January to March 2024 and telephone data collected in March 2022 and February 2024.

**Table S1:** Sociodemographic estimates from face-to-face (F2F) and telephone (Tel) data collected in 2022 and 2024 ($N_{F2F}$=4381 and $N_{Tel}$=4982).

| Sociodemographic characteristics | | Unweighted | | | | Weighted | | | |
|---|---|---|---|---|---|---|---|---|---|
| | | F2F, n | F2F, % (95% CI) | Tel, n | Tel, % (95% CI) | F2F, n | F2F, % (95% CI) | Tel, n | Tel, % (95% CI) |
| Age | 16-24 years | 602 | 13.7 (12.7-14.7) | 507 | 10.2 (9.4-11.0) | 574 | 13.1 (12.0-14.2) | 657 | 13.2 (11.9-14.4) |
| | 25-34 years | 654 | 14.9 (13.8-16.0) | 650 | 13.0 (12.1-13.9) | 727 | 16.6 (15.4-17.8) | 829 | 16.6 (15.3-18.0) |
| | 35-44 years | 687 | 15.7 (14.6-16.8) | 630 | 12.6 (11.7-13.5) | 687 | 15.7 (14.5-16.8) | 778 | 15.6 (14.3-16.9) |
| | 45-54 years | 598 | 13.6 (12.6-14.6) | 872 | 17.5 (16.4-18.6) | 717 | 16.4 (15.1-17.6) | 814 | 16.3 (15.2-17.5) |
| | 55-64 years | 699 | 16.0 (14.9-17.1) | 855 | 17.2 (16.2-18.2) | 675 | 15.4 (14.3-16.5) | 762 | 15.3 (14.1-16.4) |
| | 65+ years | 1141 | 26.0 (24.7-27.3) | 1465 | 29.4 (28.1-30.7) | 1001 | 22.9 (21.6-24.1) | 1138 | 22.9 (21.6-24.1) |
| | Refused | 0 | 0 (0-0) | 3 | 0.1 (0.0-0.2) | 0 | 0 (0-0) | 3 | 0.1 (0.0-0.1) |
| Gender | Men | 2192 | 50.1 (48.6-51.6) | 2493 | 50.3 (48.9-51.7) | 2142 | 49.0 (47.4-50.5) | 2417 | 48.8 (47.1-50.4) |
| | Women | 2181 | 49.9 (48.4-51.4) | 2440 | 49.2 (47.8-50.6) | 2231 | 51.0 (49.5-52.6) | 2516 | 50.7 (49.1-52.4) |
| | Non-binary | 0 | 0 (0-0) | 25 | 0.5 (0.3-0.7) | 0 | 0 (0-0) | 25 | 0.5 (0.3-0.7) |
| Social grade | AB | 1014 | 23.1 (21.9-24.3) | 1290 | 25.9 (24.7-27.1) | 1155 | 26.4 (24.9-27.8) | 1312 | 26.3 (24.9-27.8) |
| | C1 | 1586 | 36.2 (34.8-37.6) | 2086 | 41.9 (40.5-43.3) | 1298 | 29.6 (28.3-31.0) | 1477 | 29.6 (28.3-31.0) |
| | C2 | 782 | 17.8 (16.7-18.9) | 738 | 14.8 (13.8-15.8) | 891 | 20.3 (19.0-21.6) | 1010 | 20.3 (18.8-21.7) |
| | D | 571 | 13.0 (12.0-14.0) | 359 | 7.2 (6.5-7.9) | 637 | 14.5 (13.4-15.7) | 722 | 14.5 (13.0-16.0) |
| | E | 428 | 9.8 (8.9-10.7) | 509 | 10.2 (9.4-11.0) | 400 | 9.1 (8.3-10.0) | 460 | 9.2 (8.4-10.1) |

**Table S2:** Estimates for smoking and use of other nicotine products from face-to-face (F2F) and telephone (Tel) data collected in 2022 and 2024 ($N_{F2F}$=4381 and $N_{Tel}$=4982).

| **Smoking and use of other nicotine products** | | **Unweighted** | | | | **Weighted** | | | |
|---|---|---|---|---|---|---|---|---|---|
| | | **F2F, n** | **F2F, % (95% CI)** | **Tel, n** | **Tel, % 95% CI** | **F2F, n** | **F2F, % (95% CI)** | **Tel, n** | **Tel, % (95% CI)** |
| Tobacco use in total population | Daily cigarette smoking | 474 | 10.8 (9.9-11.7) | 478 | 9.6 (8.8-10.4) | 485 | 11.1 (10.1-12.1) | 526 | 10.6 (9.5-11.7) |
| | Non-daily cigarette smoking | 121 | 2.8 (2.3-3.3) | 145 | 2.9 (2.4-3.4) | 120 | 2.7 (2.2-3.3) | 170 | 3.4 (2.8-4.1) |
| | Non-cigarette smoking | 41 | 0.9 (0.6-1.2) | 77 | 1.5 (1.2-1.8) | 43 | 1.0 (0.7-1.3) | 87 | 1.7 (1.3-2.2) |
| | Stopped last year | 66 | 1.5 (1.1-1.9) | 94 | 1.9 (1.5-2.3) | 65 | 1.5 (1.1-1.9) | 114 | 2.3 (1.7-2.8) |
| | Stopped 1+ year ago | 785 | 17.9 (16.8-19.0) | 1178 | 23.6 (22.4-24.8) | 765 | 17.5 (16.3-18.6) | 1114 | 22.4 (21.0-23.7) |
| | Never smoked | 2883 | 65.8 (64.4-67.2) | 2953 | 59.3 (57.9-60.7) | 2892 | 66.0 (64.5-67.5) | 2912 | 58.5 (56.8-60.1) |
| | Don't know | 11 | 0.3 (0.2-0.4) | 57 | 1.1 (0.8-1.4) | 12 | 0.3 (0.1-0.4) | 59 | 1.2 (0.8-1.6) |
| Other nicotine use in total population | NRT use | 149 | 3.4 (2.9-3.9) | 135 | 2.7 (2.2-3.2) | 149 | 3.4 (2.8-4.0) | 139 | 2.8 (2.2-3.4) |
| | HTP use | 6 | 0.1 (0.0-0.2) | 12 | 0.2 (0.1-0.3) | 7 | 0.2 (0.0-0.3) | 13 | 0.3 (0.1-0.5) |
| | E-cigarette use | 259 | 5.9 (5.2-6.6) | 417 | 8.4 (7.6-9.2) | 279 | 6.4 (5.6-7.1) | 520 | 10.4 (9.3-11.5) |
| | Pouch use | 43 | 1.0 (0.7-1.3) | 19 | 0.4 (0.2-0.6) | 39 | 0.9 (0.6-1.2) | 24 | 0.5 (0.2-0.7) |
| Quitting/other nicotine use in people smoking in past year ($N_{F2F}$=702, $N_{Tel}$=794) | Tried to stop in past year | 232 | 35.7 (32.0-39.4) | 263 | 34.9 (31.5-38.3) | 241 | 36.2 (32.3-40.1) | 292 | 34.7 (30.6-38.8) |
| | Quit in past year | 66 | 9.4 (7.2-11.6) | 94 | 11.8 (9.6-14.0) | 65 | 9.1 (6.9-11.4) | 114 | 12.7 (9.8-15.6) |
| | NRT use | 123 | 17.5 (14.7-20.3) | 102 | 12.8 (10.5-15.1) | 122 | 17.1 (14.2-20.0) | 109 | 12.1 (9.5-14.8) |
| | HTP use | 5 | 0.7 (0.1-1.3) | 10 | 1.3 (0.5-2.1) | 6 | 0.8 (0.1-1.6) | 11 | 1.2 (0.2-2.3) |
| | E-cigarette use | 186 | 26.5 (23.2-29.8) | 206 | 25.9 (22.9-28.9) | 192 | 27.0 (23.5-30.5) | 263 | 29.3 (25.4-33.3) |
| | Pouch use | 19 | 2.7 (1.5-3.9) | 13 | 1.6 (0.7-2.5) | 21 | 2.9 (1.5-4.2) | 19 | 2.1 (0.8-3.3) |
| Main device type used by e-cigarette users ($N_{F2F}$=259, $N_{Tel}$=417) | Disposable | 96 | 40.3 (34.1-46.5) | 124 | 31.2 (26.6-35.8) | 98 | 38.2 (31.7-44.6) | 166 | 33.6 (28.1-39.2) |
| | Rechargeable with pre-filled cartridges | 38 | 16.0 (11.3-20.7) | 43 | 10.8 (7.8-13.8) | 38 | 14.8 (10.2-19.4) | 60 | 12.1 (8.1-16.1) |
| | Rechargeable with tank to refill | 73 | 30.7 (24.8-36.6) | 165 | 41.5 (36.7-46.3) | 84 | 32.8 (26.4-39.2) | 189 | 38.2 (32.7-43.8) |
| | Mod system | 31 | 13.0 (8.7-17.3) | 54 | 13.6 (10.2-17.0) | 37 | 14.2 (9.4-19.1) | 69 | 14.0 (9.9-18.1) |
| | Don't know | 0 | 0 (0-0) | 12 | 3.0 (1.3-4.7) | 0 | 0 (0-0) | 10 | 2.1 (0.7-3.4) |

Abbreviations: NRT, nicotine replacement therapy; HTP, heated tobacco product.

**Table S3:** Estimates for alcohol use from face-to-face (F2F) and telephone (Tel) data collected in 2022 and 2024 ($N_{F2F}$=4381 and $N_{Tel}$=4982).

| Alcohol use | | Unweighted | | | | Weighted | | | |
|---|---|---|---|---|---|---|---|---|---|
| | | **F2F, n** | **F2F, % (95% CI)** | **Tel, n** | **Tel, % (95% CI)** | **F2F, n** | **F2F, % (95% CI)** | **Tel, n** | **Tel, % (95% CI)** |
| AUDIT 1 | Never | 1408 | 32.1 (30.7-33.5) | 1208 | 24.2 (23.0-25.4) | 1363 | 31.1 (29.7-32.5) | 1246 | 25.0 (23.5-26.5) |
| | Monthly or less | 930 | 21.2 (20.0-22.4) | 1030 | 20.7 (19.6-21.8) | 954 | 21.8 (20.5-23.1) | 1072 | 21.5 (20.1-22.9) |
| | 2-4 per month | 750 | 17.1 (16.0-18.2) | 918 | 18.4 (17.3-19.5) | 782 | 17.8 (16.6-19.1) | 925 | 18.6 (17.3-19.9) |
| | 2-3 per week | 817 | 18.6 (17.4-19.8) | 1141 | 22.9 (21.7-24.1) | 823 | 18.8 (17.6-20.0) | 1082 | 21.7 (20.4-23.1) |
| | 4-5 per week | 210 | 4.8 (4.2-5.4) | 314 | 6.3 (5.6-7.0) | 203 | 4.6 (4.0-5.3) | 297 | 6.0 (5.2-6.7) |
| | 6+ per week | 229 | 5.2 (4.5-5.9) | 282 | 5.7 (5.1-6.3) | 222 | 5.1 (4.4-5.7) | 274 | 5.5 (4.8-6.2) |
| | Don't know | 4 | 0.1 (0.0-0.2) | 43 | 0.9 (0.6-1.2) | 4 | 0.1 (0.0-0.2) | 46 | 0.9 (0.6-1.3) |
| | Refused | 33 | 0.8 (0.5-1.1) | 46 | 0.9 (0.6-1.2) | 31 | 0.7 (0.5-1.0) | 40 | 0.8 (0.5-1.1) |
| AUDIT 2 ($N_{F2F}$=2973, $N_{Tel}$=3774) | 1-2 | 1425 | 48.3 (46.5-50.1) | 1715 | 45.4 (43.8-47.0) | 1439 | 48.0 (46.1-49.9) | 1667 | 44.6 (42.7-46.5) |
| | 3-4 | 824 | 27.9 (26.3-29.5) | 957 | 25.4 (24.0-26.8) | 849 | 28.3 (26.6-30.0) | 939 | 25.1 (23.5-26.8) |
| | 5-6 | 341 | 11.6 (10.4-12.8) | 425 | 11.3 (10.3-12.3) | 354 | 11.8 (10.6-13.1) | 424 | 11.3 (10.1-12.5) |
| | 7-9 | 175 | 5.9 (5.0-6.8) | 268 | 7.1 (6.3-7.9) | 172 | 5.7 (4.8-6.6) | 273 | 7.3 (6.3-8.3) |
| | 10-12 | 86 | 2.9 (2.3-3.5) | 135 | 3.6 (3-4.2.0) | 83 | 2.8 (2.2-3.4) | 139 | 3.7 (2.9-4.5) |
| | 13-15 | 37 | 1.3 (0.9-1.7) | 34 | 0.9 (0.6-1.2) | 38 | 1.3 (0.8-1.7) | 48 | 1.3 (0.8-1.8) |
| | 16+ | 37 | 1.3 (0.9-1.7) | 72 | 1.9 (1.5-2.3) | 37 | 1.2 (0.8-1.7) | 67 | 1.8 (1.3-2.3) |
| | Don't know | 11 | 0.4 (0.2-0.6) | 106 | 2.8 (2.3-3.3) | 11 | 0.4 (0.1-0.6) | 129 | 3.5 (2.6-4.3) |
| | Refused | 14 | 0.5 (0.3-0.7) | 62 | 1.6 (1.2-2.0) | 14 | 0.5 (0.2-0.8) | 51 | 1.4 (1.0-1.8) |
| AUDIT 3 ($N_{F2F}$=2973, $N_{Tel}$=3774) | Never | 1403 | 47.6 (45.8-49.4) | 1557 | 41.3 (39.7-42.9) | 1397 | 46.6 (44.7-48.5) | 1504 | 40.2 (38.4-42.1) |
| | Less than monthly | 802 | 27.2 (25.6-28.8) | 1073 | 28.4 (27.0-29.8) | 851 | 28.4 (26.7-30.1) | 1099 | 29.4 (27.7-31.2) |
| | Monthly | 388 | 13.2 (12.0-14.4) | 556 | 14.7 (13.6-15.8) | 393 | 13.1 (11.8-14.4) | 558 | 14.9 (13.5-16.3) |
| | Weekly | 300 | 10.2 (9.1-11.3) | 414 | 11.0 (10.0-12.0) | 299 | 10.0 (8.8-11.1) | 412 | 11.0 (9.8-12.3) |
| | Daily or almost daily | 35 | 1.2 (0.8-1.6) | 71 | 1.9 (1.5-2.3) | 35 | 1.2 (0.8-1.6) | 73 | 1.9 (1.4-2.5) |
| | Don't know | 7 | 0.2 (0.0-0.4) | 51 | 1.4 (1.0-1.8) | 8 | 0.3 (0.1-0.5) | 49 | 1.3 (0.9-1.8) |
| | Refused | 15 | 0.5 (0.2-0.8) | 52 | 1.4 (1.0-1.8) | 15 | 0.5 (0.2-0.8) | 41 | 1.1 (0.7-1.5) |
| AUDIT-C ≥5 | | 1090 | 25.2 (23.9-26.5) | 1455 | 30.6 (29.3-31.9) | 1095 | 25.3 (23.9-26.7) | 1447 | 30.4 (28.9-32.0) |

AUDIT 1: how often drinking; AUDIT 2: how many standard drinks on a typical drinking day; AUDIT 3: how often ≥6 standard drinks on one occasion; AUDIT-C ≥5: increasing and higher risk drinking.

### 3. Comparing 2024 data using telephone data from February 2024

This section includes face-to-face data collected between January and March 2024 and telephone data collected in February 2024.

**Table S4**: Missing values during face-to-face (F2F) and telephone (Tel) data collected in 2024 (F2F: Jan-Mar; Tel: Feb; $N_{F2F}$=2317, $N_{Tel}$=2375).

| **Variable** | **F2F missing values** | | **Tel missing values** | |
|---|---|---|---|---|
| | **n** | **% (95% CI)** | **n** | **% (95% CI)** |
| Age | 0 | 0 (0-0) | 0 | 0 (0-0) |
| Gender | 8 | 0.3 (0.1-0.5) | 24 | 1.0 (0.6-1.4) |
| Social grade | 0 | 0 (0-0) | 0 | 0 (0-0) |
| Smoking status | 0 | 0 (0-0) | 0 | 0 (0-0) |
| AUDIT 1 | 0 | 0 (0-0) | 0 | 0 (0-0) |
| AUDIT 2 and 3 ($N_{F2F}$=1541, $N_{Tel}$=1766)[1] | 1 | 0.1 (0.0-0.3) | 0 | 0 (0-0) |
| AUDIT-C ≥5 | 18 | 0.8 (0.4-1.2) | 83 | 3.5 (2.8-4.2) |
| Quit attempt in past year ($N_{F2F}$=317, $N_{Tel}$=364)[2] | 20 | 6.3 (3.6-9.0) | 13 | 3.6 (1.7-5.5) |
| E-cigarette device used ($N_{F2F}$=139, $N_{Tel}$=241)[3] | 13 | 9.4 (4.5-14.3) | 2 | 0.8 (0.0-1.9) |

[1] only assessed for people who reported drinking alcohol for AUDIT 1 question.

[2] only assessed for people who reported smoking in the past year.

[3] only assessed for people who reported using e-cigarettes.

**Table S5:** Sociodemographic estimates from face-to-face (F2F) and telephone (Tel) data collected in 2024 (F2F: Jan-Mar; Tel: Feb; $N_{F2F}$=2317 and $N_{Tel}$=2375).

| **Sociodemographic characteristics** | | **Unweighted** | | | | **Weighted** | | | |
|---|---|---|---|---|---|---|---|---|---|
| | | **F2F, n** | **F2F, % (95% CI)** | **Tel, n** | **Tel, % (95% CI)** | **F2F, n** | **F2F, % (95% CI)** | **Tel, n** | **Tel, % (95% CI)** |
| Age | 16-24 years | 257 | 11.1 (9.8-12.4) | 234 | 9.9 (8.7-11.1) | 298 | 12.9 (11.3-14.4) | 306 | 12.9 (11.2-14.6) |
| | 25-34 years | 333 | 14.4 (13.0-15.8) | 277 | 11.7 (10.4-13.0) | 384 | 16.6 (14.9-18.3) | 392 | 16.5 (14.6-18.4) |
| | 35-44 years | 375 | 16.2 (14.7-17.7) | 326 | 13.7 (12.3-15.1) | 367 | 15.8 (14.3-17.4) | 375 | 15.8 (14.1-17.5) |
| | 45-54 years | 314 | 13.6 (12.2-15.0) | 403 | 17.0 (15.5-18.5) | 374 | 16.2 (14.5-17.8) | 383 | 16.1 (14.5-17.7) |
| | 55-64 years | 361 | 15.6 (14.1-17.1) | 409 | 17.2 (15.7-18.7) | 364 | 15.7 (14.1-17.2) | 373 | 15.7 (14.1-17.3) |
| | 65+ years | 677 | 29.2 (27.3-31.1) | 725 | 30.5 (28.6-32.4) | 530 | 22.9 (21.2-24.5) | 546 | 23.0 (21.2-24.7) |
| | Refused | 0 | 0 (0-0) | 1 | 0.0 (0.0-0.1) | 0 | 0 (0-0) | 1 | 0.0 (0.0-0.1) |
| Gender | Men | 1167 | 50.5 (48.5-52.5) | 1233 | 52.4 (50.4-54.4) | 1132 | 49.0 (46.9-51.2) | 1153 | 49.0 (46.7-51.4) |
| | Women | 1142 | 49.5 (47.5-51.5) | 1118 | 47.6 (45.6-49.6) | 1177 | 51.0 (48.8-53.1) | 1198 | 51.0 (48.6-53.3) |
| | Non-binary | 0 | 0 (0-0) | 0 | 0 (0-0) | 0 | 0 (0-0) | 0 | 0 (0-0) |
| Social grade | AB | 606 | 26.2 (24.4-28.0) | 687 | 28.9 (27.1-30.7) | 610 | 26.3 (24.4-28.2) | 624 | 26.3 (24.3-28.2) |
| | C1 | 782 | 33.8 (31.9-35.7) | 936 | 39.4 (37.4-41.4) | 688 | 29.7 (27.8-31.6) | 704 | 29.7 (27.7-31.6) |
| | C2 | 411 | 17.7 (16.1-19.3) | 342 | 14.4 (13.0-15.8) | 471 | 20.3 (18.5-22.1) | 482 | 20.3 (18.3-22.3) |
| | D | 302 | 13.0 (11.6-14.4) | 178 | 7.5 (6.4-8.6) | 336 | 14.5 (12.9-16.1) | 344 | 14.5 (12.4-16.6) |
| | E | 216 | 9.3 (8.1-10.5) | 232 | 9.8 (8.6-11.0) | 212 | 9.1 (7.9-10.3) | 221 | 9.3 (8.0-10.5) |

**Table S6:** Estimates for smoking and use of other nicotine products from face-to-face (F2F) and telephone (Tel) data collected in 2024 (F2F: Jan-Mar; Tel: Feb; $N_{F2F}$=2317 and $N_{Tel}$=2375).

| **Smoking and use of other nicotine products** | | **Unweighted** | | | | **Weighted** | | | |
|---|---|---|---|---|---|---|---|---|---|
| | | **F2F, n** | **F2F, % (95% CI)** | **Tel, n** | **Tel, % 95% CI** | **F2F, n** | **F2F, % (95% CI)** | **Tel, n** | **Tel, % (95% CI)** |
| Tobacco use in total population | Daily cigarette smoking | 212 | 9.1 (7.9-10.3) | 208 | 8.8 (7.7-9.9) | 230 | 9.9 (8.6-11.2) | 233 | 9.8 (8.4-11.3) |
| | Non-daily cigarette smoking | 47 | 2.0 (1.4-2.6) | 75 | 3.2 (2.5-3.9) | 47 | 2.0 (1.4-2.7) | 87 | 3.7 (2.8-4.6) |
| | Non-cigarette smoking | 25 | 1.1 (0.7-1.5) | 28 | 1.2 (0.8-1.6) | 26 | 1.1 (0.7-1.6) | 34 | 1.4 (0.8-2.0) |
| | Stopped last year | 33 | 1.4 (0.9-1.9) | 53 | 2.2 (1.6-2.8) | 34 | 1.5 (0.9-2.0) | 62 | 2.6 (1.8-3.4) |
| | Stopped 1+ year ago | 445 | 19.2 (17.6-20.8) | 582 | 24.5 (22.8-26.2) | 423 | 18.2 (16.6-19.9) | 563 | 23.7 (21.8-25.6) |
| | Never smoked | 1551 | 66.9 (65.0-68.8) | 1411 | 59.4 (57.4-61.4) | 1553 | 67.0 (65.0-69.1) | 1376 | 57.9 (55.6-60.2) |
| | Don’t know | 4 | 0.2 (0.0-0.4) | 18 | 0.8 (0.5-1.1) | 3 | 0.1 (0.0-0.3) | 20 | 0.9 (0.4-1.3) |
| Other nicotine use in total population | NRT use | 69 | 3.0 (2.3-3.7) | 65 | 2.7 (2.0-3.4) | 69 | 3.0 (2.3-3.7) | 70 | 3.0 (2.2-3.7) |
| | HTP use | 2 | 0.1 (0.0-0.2) | 7 | 0.3 (0.1-0.5) | 2 | 0.1 (0.0-0.2) | 6 | 0.3 (0.1-0.5) |
| | E-cigarette use | 139 | 6.0 (5.0-7.0) | 241 | 10.1 (8.9-11.3) | 164 | 7.1 (5.9-8.2) | 299 | 12.6 (10.9-14.3) |
| | Pouch use | 32 | 1.4 (0.9-1.9) | 10 | 0.4 (0.1-0.7) | 27 | 1.2 (0.8-1.6) | 12 | 0.5 (0.1-0.9) |
| Quitting/other nicotine use in people smoking in past year ($N_{F2F}$=317, $N_{Tel}$=364) | Tried to stop in past year | 113 | 38.0 (32.5-43.5) | 118 | 33.6 (28.7-38.5) | 121 | 38.0 (32.2-43.8) | 132 | 32.7 (27.1-38.4) |
| | Quit in past year | 33 | 10.4 (7.0-13.8) | 53 | 14.6 (11.0-18.2) | 34 | 10.0 (6.6-13.5) | 62 | 15.0 (10.8-19.2) |
| | NRT use | 43 | 13.6 (9.8-17.4) | 49 | 13.5 (10.0-17.0) | 43 | 12.7 (9.0-16.3) | 55 | 13.1 (9.3-16.9) |
| | HTP use | 1 | 0.3 (0.0-0.9) | 5 | 1.4 (0.2-2.6) | 1 | 0.4 (0.0-1.0) | 4 | 1.0 (0.1-1.9) |
| | E-cigarette use | 66 | 20.8 (16.3-25.3) | 105 | 28.8 (24.1-33.5) | 78 | 23.0 (18.0-28.0) | 136 | 32.6 (26.9-38.3) |
| | Pouch use | 8 | 2.5 (0.8-4.2) | 9 | 2.5 (0.9-4.1) | 9 | 2.6 (0.7-4.4) | 11 | 2.6 (0.7-4.6) |
| Main device type used by e-cigarette users ($N_{F2F}$=139, $N_{Tel}$=241) | Disposable | 47 | 37.3 (28.9-45.7) | 92 | 38.5 (32.3-44.7) | 55 | 36.8 (28.0-45.5) | 125 | 42.0 (34.8-49.2) |
| | Rechargeable with pre-filled cartridges | 17 | 13.5 (7.5-19.5) | 19 | 7.9 (4.5-11.3) | 17 | 11.5 (6.0-17.1) | 27 | 9.1 (4.8-13.4) |
| | Rechargeable with tank to refill | 38 | 30.2 (22.2-38.2) | 96 | 40.2 (34.0-46.4) | 48 | 32.0 (23.3-40.6) | 107 | 36.0 (29.3-42.8) |
| | Mod system | 24 | 19.0 (12.1-25.9) | 26 | 10.9 (7.0-14.8) | 30 | 19.8 (12.3-27.2) | 33 | 11.2 (6.6-15.9) |
| | Don’t know | 0 | 0 (0-0) | 6 | 2.5 (0.5-4.5) | 0 | 0 (0-0) | 5 | 1.7 (0.2-3.1) |

Abbreviations: NRT, nicotine replacement therapy; HTP, heated tobacco product.

**Table S7:** Estimates for alcohol use from face-to-face (F2F) and telephone (Tel) data collected in 2024 (F2F: Jan-Mar; Tel: Feb; $N_{F2F}$=2317 and $N_{Tel}$=2375).

| **Alcohol use** | | **Unweighted** | | | | **Weighted** | | | |
|---|---|---|---|---|---|---|---|---|---|
| | | **F2F, n** | **F2F, % (95% CI)** | **Tel, n** | **Tel, % (95% CI)** | **F2F, n** | **F2F, % (95% CI)** | **Tel, n** | **Tel, % (95% CI)** |
| AUDIT 1 | Never | 775 | 33.4 (31.5-35.3) | 609 | 25.6 (23.8-27.4) | 733 | 31.6 (29.7-33.6) | 628 | 26.4 (24.4-28.5) |
| | Monthly or less | 522 | 22.5 (20.8-24.2) | 483 | 20.3 (18.7-21.9) | 542 | 23.4 (21.5-25.2) | 521 | 21.9 (20.0-23.9) |
| | 2-4 per month | 408 | 17.6 (16.0-19.2) | 433 | 18.2 (16.6-19.8) | 436 | 18.8 (17.1-20.5) | 435 | 18.3 (16.5-20.1) |
| | 2-3 per week | 390 | 16.8 (15.3-18.3) | 530 | 22.3 (20.6-24.0) | 397 | 17.1 (15.5-18.8) | 507 | 21.3 (19.5-23.2) |
| | 4-5 per week | 103 | 4.4 (3.6-5.2) | 154 | 6.5 (5.5-7.5) | 98 | 4.2 (3.4-5.1) | 130 | 5.5 (4.5-6.5) |
| | 6+ per week | 108 | 4.7 (3.8-5.6) | 133 | 5.6 (4.7-6.5) | 100 | 4.3 (3.5-5.1) | 122 | 5.1 (4.2-6.1) |
| | Don't know | 2 | 0.1 (0.0-0.2) | 15 | 0.6 (0.3-0.9) | 2 | 0.1 (0.0-0.2) | 14 | 0.6 (0.2-0.9) |
| | Refused | 9 | 0.4 (0.1-0.7) | 18 | 0.8 (0.5-1.1) | 9 | 0.4 (0.1-0.7) | 18 | 0.8 (0.4-1.2) |
| AUDIT 2 ($N_{F2F}$=1541, $N_{Tel}$=1766) | 1-2 | 739 | 48.0 (45.5-50.5) | 810 | 45.9 (43.6-48.2) | 728 | 46 (43.3-48.6) | 765 | 43.8 (41.1-46.4) |
| | 3-4 | 446 | 28.9 (26.6-31.2) | 466 | 26.4 (24.3-28.5) | 471 | 29.8 (27.3-32.2) | 462 | 26.4 (24.1-28.8) |
| | 5-6 | 172 | 11.2 (9.6-12.8) | 202 | 11.4 (9.9-12.9) | 189 | 11.9 (10.2-13.7) | 208 | 11.9 (10.2-13.7) |
| | 7-9 | 90 | 5.8 (4.6-7.0) | 125 | 7.1 (5.9-8.3) | 96 | 6.0 (4.8-7.3) | 133 | 7.6 (6.2-9.1) |
| | 10-12 | 44 | 2.9 (2.1-3.7) | 53 | 3.0 (2.2-3.8) | 46 | 2.9 (2.0-3.8) | 55 | 3.2 (2.2-4.2) |
| | 13-15 | 17 | 1.1 (0.6-1.6) | 18 | 1.0 (0.5-1.5) | 17 | 1.1 (0.5-1.6) | 27 | 1.5 (0.8-2.3) |
| | 16+ | 17 | 1.1 (0.6-1.6) | 33 | 1.9 (1.3-2.5) | 20 | 1.2 (0.6-1.9) | 33 | 1.9 (1.1-2.6) |
| | Don't know | 5 | 0.3 (0.0-0.6) | 38 | 2.2 (1.5-2.9) | 6 | 0.4 (0.0-0.7) | 44 | 2.5 (1.6-3.5) |
| | Refused | 11 | 0.7 (0.3-1.1) | 21 | 1.2 (0.7-1.7) | 12 | 0.8 (0.3-1.3) | 20 | 1.1 (0.6-1.7) |
| AUDIT 3 ($N_{F2F}$=1541, $N_{Tel}$=1766) | Never | 741 | 48.1 (45.6-50.6) | 728 | 41.2 (38.9-43.5) | 716 | 45.2 (42.6-47.8) | 683 | 39.1 (36.5-41.7) |
| | Less than monthly | 420 | 27.3 (25.1-29.5) | 511 | 28.9 (26.8-31.0) | 460 | 29.0 (26.6-31.5) | 539 | 30.9 (28.3-33.4) |
| | Monthly | 195 | 12.7 (11.0-14.4) | 266 | 15.1 (13.4-16.8) | 215 | 13.6 (11.7-15.4) | 267 | 15.3 (13.3-17.2) |
| | Weekly | 151 | 9.8 (8.3-11.3) | 187 | 10.6 (9.2-12.0) | 156 | 9.9 (8.3-11.5) | 184 | 10.5 (8.9-12.2) |
| | Daily or almost daily | 19 | 1.2 (0.6-1.8) | 33 | 1.9 (1.3-2.5) | 20 | 1.2 (0.7-1.8) | 36 | 2.1 (1.2-2.9) |
| | Don't know | 4 | 0.3 (0.0-0.6) | 21 | 1.2 (0.7-1.7) | 4 | 0.3 (0.0-0.5) | 19 | 1.1 (0.5-1.7) |
| | Refused | 11 | 0.7 (0.3-1.1) | 20 | 1.1 (0.6-1.6) | 12 | 0.8 (0.3-1.3) | 19 | 1.1 (0.5-1.6) |
| AUDIT-C ≥5 | | 540 | 23.5 (21.8-25.2) | 680 | 29.7 (27.8-31.6) | 567 | 24.7 (22.8-26.6) | 675 | 29.5 (27.3-31.6) |

AUDIT 1: how often drinking; AUDIT 2: how many standard drinks on a typical drinking day; AUDIT 3: how often ≥6 standard drinks on one occasion; AUDIT-C ≥5: increasing and higher risk drinking.

### 4. Comparing 2024 data using telephone data from January to March 2024

This section includes face-to-face data collected between January and March 2024 and telephone data collected between January and March 2024.

Reweighting applied to telephone data:

- for January in which 21% were collected, weights were multiplied by approximately 0.27 [21%/(65%+14%)];
- for February in which 65% were collected, weights were multiplied by approximately 1.86 [65%/(21%+14%)];
- for March in which 14% were collected, weights were multiplied by approximately 0.16 [14%/(21%+65%)]).

**Table S8:** Sociodemographic estimates from face-to-face (F2F) and telephone (Tel) data collected in Jan-Mar 2024 ($N_{F2F}$=2317 and $N_{Tel\text{-}3\text{-}months}$=7157).

| **Sociodemographic characteristics** | | **Unweighted** | | | | **Weighted** | | | |
|---|---|---|---|---|---|---|---|---|---|
| | | **F2F, n** | **F2F, % (95% CI)** | **Tel, n** | **Tel, % (95% CI)** | **F2F, n** | **F2F, % (95% CI)** | **Tel, n** | **Tel, % (95% CI)** |
| Age | 16-24 years | 257 | 11.1 (9.8-12.4) | 732 | 10.2 (9.5-10.9) | 298 | 12.9 (11.3-14.4) | 921 | 12.9 (11.9-13.9) |
| | 25-34 years | 333 | 14.4 (13.0-15.8) | 862 | 12.0 (11.2-12.8) | 384 | 16.6 (14.9-18.3) | 1189 | 16.6 (15.5-17.7) |
| | 35-44 years | 375 | 16.2 (14.7-17.7) | 893 | 12.5 (11.7-13.3) | 367 | 15.8 (14.3-17.4) | 1132 | 15.8 (14.8-16.9) |
| | 45-54 years | 314 | 13.6 (12.2-15.0) | 1151 | 16.1 (15.2-17.0) | 374 | 16.2 (14.5-17.8) | 1149 | 16.1 (15.1-17.0) |
| | 55-64 years | 361 | 15.6 (14.1-17.1) | 1313 | 18.3 (17.4-19.2) | 364 | 15.7 (14.1-17.2) | 1121 | 15.7 (14.7-16.6) |
| | 65+ years | 677 | 29.2 (27.3-31.1) | 2200 | 30.7 (29.6-31.8) | 530 | 22.9 (21.2-24.5) | 1639 | 22.9 (21.9-23.9) |
| | Refused | 0 | 0 (0-0) | 6 | 0.1 (0.0-0.2) | 0 | 0 (0-0) | 6 | 0.1 (0.0-0.2) |
| Gender | Men | 1167 | 50.5 (48.5-52.5) | 3566 | 50.4 (49.2-51.6) | 1132 | 49.0 (46.9-51.2) | 3462 | 49.0 (47.6-50.3) |
| | Women | 1142 | 49.5 (47.5-51.5) | 3495 | 49.4 (48.2-50.6) | 1177 | 51.0 (48.8-53.1) | 3599 | 50.9 (49.5-52.3) |
| | Non-binary | 0 | 0 (0-0) | 8 | 0.1 (0.0-0.2) | 0 | 0 (0-0) | 8 | 0.1 (0.0-0.2) |
| Social grade | AB | 606 | 26.2 (24.4-28.0) | 2142 | 29.9 (28.8-31.0) | 610 | 26.3 (24.4-28.2) | 1878 | 26.2 (25.1-27.3) |
| | C1 | 782 | 33.8 (31.9-35.7) | 2825 | 39.5 (38.4-40.6) | 688 | 29.7 (27.8-31.6) | 2128 | 29.7 (28.6-30.8) |
| | C2 | 411 | 17.7 (16.1-19.3) | 969 | 13.5 (12.7-14.3) | 471 | 20.3 (18.5-22.1) | 1449 | 20.3 (19.0-21.5) |
| | D | 302 | 13.0 (11.6-14.4) | 512 | 7.2 (6.6-7.8) | 336 | 14.5 (12.9-16.1) | 1036 | 14.5 (13.2-15.7) |
| | E | 216 | 9.3 (8.1-10.5) | 709 | 9.9 (9.2-10.6) | 212 | 9.1 (7.9-10.3) | 665 | 9.3 (8.6-10.0) |

**Table S9**: Estimates for smoking and use of other nicotine products from face-to-face (F2F) and telephone (Tel) data collected in Jan-Mar 2024 ($N_{F2F}$=2317 and $N_{Tel\text{-}3\text{-}months}$=7157).

| **Smoking and use of other nicotine products** | | **Unweighted** | | | | **Weighted** | | | |
|---|---|---|---|---|---|---|---|---|---|
| | | **F2F, n** | **F2F, % (95% CI)** | **Tel, n** | **Tel, % 95% CI** | **F2F, n** | **F2F, % (95% CI)** | **Tel, n** | **Tel, % (95% CI)** |
| Tobacco use in total population | Daily cigarette smoking | 212 | 9.1 (7.9-10.3) | 628 | 8.8 (8.1-9.5) | 230 | 9.9 (8.6-11.2) | 750 | 10.5 (9.6-11.4) |
| | Non-daily cigarette smoking | 47 | 2.0 (1.4-2.6) | 204 | 2.9 (2.5-3.3) | 47 | 2.0 (1.4-2.7) | 242 | 3.4 (2.9-3.9) |
| | Non-cigarette smoking | 25 | 1.1 (0.7-1.5) | 112 | 1.6 (1.3-1.9) | 26 | 1.1 (0.7-1.6) | 121 | 1.7 (1.3-2.1) |
| | Stopped last year | 33 | 1.4 (0.9-1.9) | 167 | 2.3 (2.0-2.6) | 34 | 1.5 (0.9-2.0) | 202 | 2.8 (2.3-3.3) |
| | Stopped 1+ year ago | 445 | 19.2 (17.6-20.8) | 1771 | 24.7 (23.7-25.7) | 423 | 18.2 (16.6-19.9) | 1667 | 23.3 (22.2-24.4) |
| | Never smoked | 1551 | 66.9 (65.0-68.8) | 4224 | 59.0 (57.9-60.1) | 1553 | 67.0 (65.0-69.1) | 4119 | 57.5 (56.2-58.9) |
| | Don't know | 4 | 0.2 (0.0-0.4) | 51 | 0.7 (0.5-0.9) | 3 | 0.1 (0.0-0.3) | 56 | 0.8 (0.5-1.0) |
| Other nicotine use in total population | NRT use | 69 | 3.0 (2.3-3.7) | 189 | 2.6 (2.2-3.0) | 69 | 3.0 (2.3-3.7) | 206 | 2.9 (2.4-3.4) |
| | HTP use | 2 | 0.1 (0.0-0.2) | 12 | 0.2 (0.1-0.3) | 2 | 0.1 (0.0-0.2) | 13 | 0.2 (0.1-0.3) |
| | E-cigarette use | 139 | 6.0 (5.0-7.0) | 676 | 9.4 (8.7-10.1) | 164 | 7.1 (5.9-8.2) | 847 | 11.8 (10.9-12.8) |
| | Pouch use | 32 | 1.4 (0.9-1.9) | 39 | 0.5 (0.3-0.7) | 27 | 1.2 (0.8-1.6) | 49 | 0.7 (0.4-0.9) |
| Quitting/other nicotine use in people smoking in past year ($N_{F2F}$=317, $N_{Tel\text{-}3\text{-}months}$=1111) | Tried to stop in past year | 113 | 38.0 (32.5-43.5) | 362 | 33.9 (31.1-36.7) | 121 | 38.0 (32.2-43.8) | 434 | 34.3 (30.9-37.7) |
| | Quit in past year | 33 | 10.4 (7.0-13.8) | 167 | 15.0 (12.9-17.1) | 34 | 10.0 (6.6-13.5) | 202 | 15.4 (12.9-17.8) |
| | NRT use | 43 | 13.6 (9.8-17.4) | 136 | 12.2 (10.3-14.1) | 43 | 12.7 (9.0-16.3) | 156 | 11.9 (9.6-14.1) |
| | HTP use | 1 | 0.3 (0.0-0.9) | 8 | 0.7 (0.2-1.2) | 1 | 0.4 (0.0-1.0) | 8 | 0.6 (0.2-1.1) |
| | E-cigarette use | 66 | 20.8 (16.3-25.3) | 312 | 28.1 (25.5-30.7) | 78 | 23.0 (18.0-28.0) | 410 | 31.2 (27.9-34.5) |
| | Pouch use | 8 | 2.5 (0.8-4.2) | 28 | 2.5 (1.6-3.4) | 9 | 2.6 (0.7-4.4) | 36 | 2.7 (1.6-3.9) |
| Main device type used by e-cigarette users ($N_{F2F}$=139, $N_{Tel\text{-}3\text{-}months}$=676) | Disposable | 47 | 37.3 (28.9-45.7) | 241 | 36.0 (32.4-39.6) | 55 | 36.8 (28.0-45.5) | 324 | 38.5 (34.2-42.9) |
| | Rechargeable with pre-filled cartridges | 17 | 13.5 (7.5-19.5) | 71 | 10.6 (8.3-12.9) | 17 | 11.5 (6.0-17.1) | 92 | 10.9 (8.1-13.7) |
| | Rechargeable with tank to refill | 38 | 30.2 (22.2-38.2) | 261 | 39.0 (35.3-42.7) | 48 | 32.0 (23.3-40.6) | 308 | 36.7 (32.5-40.8) |
| | Mod system | 24 | 19.0 (12.1-25.9) | 89 | 13.3 (10.7-15.9) | 30 | 19.8 (12.3-27.2) | 110 | 13.1 (10.1-16.1) |
| | Don't know | 0 | 0 (0-0) | 8 | 1.2 (0.4-2.0) | 0 | 0 (0-0) | 7 | 0.9 (0.2-1.5) |

Abbreviations: NRT, nicotine replacement therapy; HTP, heated tobacco product.

**Table S10:** Estimates for alcohol use from face-to-face (F2F) and telephone (Tel) data collected in Jan-Mar 2024 ($N_{F2F}$=2317 and $N_{Tel\text{-}3\text{-}months}$=7157).

| **Alcohol use** | | **Unweighted** | | | | **Weighted** | | | |
|---|---|---|---|---|---|---|---|---|---|
| | | **F2F, n** | **F2F, % (95% CI)** | **Tel, n** | **Tel, % (95% CI)** | **F2F, n** | **F2F, % (95% CI)** | **Tel, n** | **Tel, % (95% CI)** |
| AUDIT 1 | Never | 775 | 33.4 (31.5-35.3) | 1747 | 24.4 (23.4-25.4) | 733 | 31.6 (29.7-33.6) | 1829 | 25.6 (24.4-26.8) |
| | Monthly or less | 522 | 22.5 (20.8-24.2) | 1534 | 21.4 (20.4-22.4) | 542 | 23.4 (21.5-25.2) | 1646 | 23.0 (21.8-24.2) |
| | 2-4 per month | 408 | 17.6 (16.0-19.2) | 1314 | 18.4 (17.5-19.3) | 436 | 18.8 (17.1-20.5) | 1331 | 18.6 (17.5-19.7) |
| | 2-3 per week | 390 | 16.8 (15.3-18.3) | 1597 | 22.3 (21.3-23.3) | 397 | 17.1 (15.5-18.8) | 1514 | 21.2 (20.1-22.2) |
| | 4-5 per week | 103 | 4.4 (3.6-5.2) | 467 | 6.5 (5.9-7.1) | 98 | 4.2 (3.4-5.1) | 398 | 5.6 (5.0-6.1) |
| | 6+ per week | 108 | 4.7 (3.8-5.6) | 420 | 5.9 (5.4-6.4) | 100 | 4.3 (3.5-5.1) | 363 | 5.1 (4.5-5.6) |
| | Don't know | 2 | 0.1 (0-0.2) | 41 | 0.6 (0.4-0.8) | 2 | 0.1 (0.0-0.2) | 40 | 0.6 (0.4-0.8) |
| | Refused | 9 | 0.4 (0.1-0.7) | 37 | 0.5 (0.3-0.7) | 9 | 0.4 (0.1-0.7) | 36 | 0.5 (0.3-0.7) |
| AUDIT 2 ($N_{F2F}$=1541, $N_{Tel\text{-}3\text{-}months}$= 5410) | 1-2 | 739 | 48 (45.5-50.5) | 2520 | 46.6 (45.3-47.9) | 728 | 46 (43.3-48.6) | 2357 | 44.2 (42.7-45.8) |
| | 3-4 | 446 | 28.9 (26.6-31.2) | 1420 | 26.2 (25.0-27.4) | 471 | 29.8 (27.3-32.2) | 1420 | 26.7 (25.3-28) |
| | 5-6 | 172 | 11.2 (9.6-12.8) | 619 | 11.4 (10.6-12.2) | 189 | 11.9 (10.2-13.7) | 632 | 11.9 (10.9-12.9) |
| | 7-9 | 90 | 5.8 (4.6-7) | 356 | 6.6 (5.9-7.3) | 96 | 6.0 (4.8-7.3) | 389 | 7.3 (6.4-8.2) |
| | 10-12 | 44 | 2.9 (2.1-3.7) | 193 | 3.6 (3.1-4.1) | 46 | 2.9 (2.0-3.8) | 207 | 3.9 (3.3-4.5) |
| | 13-15 | 17 | 1.1 (0.6-1.6) | 48 | 0.9 (0.7-1.1) | 17 | 1.1 (0.5-1.6) | 63 | 1.2 (0.8-1.6) |
| | 16+ | 17 | 1.1 (0.6-1.6) | 99 | 1.8 (1.4-2.2) | 20 | 1.2 (0.6-1.9) | 107 | 2.0 (1.5-2.5) |
| | Don't know | 5 | 0.3 (0-0.6) | 105 | 1.9 (1.5-2.3) | 6 | 0.4 (0.0-0.7) | 108 | 2.0 (1.6-2.5) |
| | Refused | 11 | 0.7 (0.3-1.1) | 50 | 0.9 (0.6-1.2) | 12 | 0.8 (0.3-1.3) | 45 | 0.8 (0.6-1.1) |
| AUDIT 3 ($N_{F2F}$=1541, $N_{Tel\text{-}3\text{-}months}$= 5410) | Never | 741 | 48.1 (45.6-50.6) | 2260 | 41.8 (40.5-43.1) | 716 | 45.2 (42.6-47.8) | 2077 | 39.0 (37.5-40.5) |
| | Less than monthly | 420 | 27.3 (25.1-29.5) | 1636 | 30.2 (29.0-31.4) | 460 | 29.0 (26.6-31.5) | 1730 | 32.5 (31.0-34.0) |
| | Monthly | 195 | 12.7 (11.0-14.4) | 792 | 14.6 (13.7-15.5) | 215 | 13.6 (11.7-15.4) | 803 | 15.1 (13.9-16.2) |
| | Weekly | 151 | 9.8 (8.3-11.3) | 531 | 9.8 (9.0-10.6) | 156 | 9.9 (8.3-11.5) | 529 | 9.9 (9.0-10.9) |
| | Daily or almost daily | 19 | 1.2 (0.6-1.8) | 109 | 2.0 (1.6-2.4) | 20 | 1.2 (0.7-1.8) | 107 | 2.0 (1.6-2.4) |
| | Don't know | 4 | 0.3 (0.0-0.6) | 44 | 0.8 (0.6-1.0) | 4 | 0.3 (0.0-0.5) | 45 | 0.8 (0.5-1.1) |
| | Refused | 11 | 0.7 (0.3-1.1) | 38 | 0.7 (0.5-0.9) | 12 | 0.8 (0.3-1.3) | 36 | 0.7 (0.4-0.9) |
| AUDIT-C ≥5 | | 540 | 23.5 (21.8-25.2) | 2089 | 30.1 (29.0-31.2) | 567 | 24.7 (22.8-26.6) | 2072 | 29.8 (28.6-31.1) |

AUDIT 1: how often drinking; Audit 2: how many standard drinks on a typical drinking day; AUDIT 3: how often ≥6 standard drinks on one occasion; AUDIT-C ≥5: increasing and higher risk drinking.

### 5. Exploring differences in e-cigarette use

This section further explores differences in e-cigarette use by comparing e-cigarette use in the total population stratified by age and (given e-cigarette use was assessed within several different survey questions) by counting the number and proportion of participants who reported e-cigarette use in response to the different survey questions that contributed to the derived variable "e-cigarette use in total population".

**Table S11:** Estimates for e-cigarette use in the total population stratified by age from face-to-face (F2F) and telephone (Tel) data collected in 2022 and 2024 ($N_{F2F}$=4381 and $N_{Tel}$=4982).

| Age group | Unweighted | | | | Weighted | | | |
|---|---|---|---|---|---|---|---|---|
| | F2F, n | F2F, % (95% CI) | Tel, n | Tel, % (95% CI) | F2F, n | F2F, % (95% CI) | Tel, n | Tel, % (95% CI) |
| 16-24 | 59 | 9.8 (7.1-12.5) | 99 | 19.5 (17.3-21.7) | 58 | 10.1 (7.5-12.7) | 141 | 21.5 (17.2- 25.8) |
| 25-34 | 58 | 8.9 (8.0-9.8) | 98 | 15.1(14.0-16.2) | 65 | 9.0 (6.7-11.3) | 148 | 17.8 (14.1-21.5) |
| 35-44 | 41 | 6.0 (3.3-8.7) | 78 | 12.4 (10.1-14.7) | 46 | 6.7 (4.6-8.7) | 101 | 13.0 (9.9-16.0) |
| 45-54 | 40 | 6.7 (5.9-7.5) | 57 | 6.5 (5.5-7.5) | 52 | 7.2 (5.0-9.4) | 53 | 6.6 (4.7-8.4) |
| 55-64 | 40 | 5.7 (3.0-8.4) | 44 | 5.1 (2.8-7.4) | 39 | 5.7 (3.9-7.5) | 38 | 5.0 (3.3-6.7) |
| 65+ | 21 | 1.8 (1.1-2.5) | 40 | 2.7 (1.8-3.6) | 19 | 1.9 (1.1-2.8) | 38 | 3.3 (2.1-4.4) |

**Table S12:** Unweighted number and proportion of positive responses to each variable that contributed to the derived variable "current e-cigarette use in total population", based on face-to-face (F2F) and telephone (Tel) data collected in 2022 and 2024 ($N_{F2F}$=4381 and $N_{Tel}$=4982). Responses not mutually exclusive.

| Survey variable with e-cigarette or Juul as response option | F2F | | | Tel | | |
|---|---|---|---|---|---|---|
| | N | n | % (95% CI) | N | n | % (95% CI) |
| Using e-cigarette to stop smoking, cut down, or any other reason[1] | 702 | 140 | 19.9 (16.9-22.9) | 794 | 150 | 18.9 (16.2-21.6) |
| Using Juul to stop smoking, cut down, or any other reason[1] | 702 | 12 | 1.7 (0.7-2.7) | 794 | 5 | 0.6 (0.0-1.2) |
| Using e-cigarette to help cut down amount smoked[2] | 277 | 82 | 29.6 (24.2-35.0) | 362 | 101 | 27.9 (23.3-32.5) |
| Using Juul to help cut down amount smoked[2] | 277 | 7 | 2.5 (0.7-4.3) | 362 | 6 | 1.7 (0.4-3.0) |
| Regularly using e-cigarette when not allowed to smoke[2] | 636 | 118 | 18.6 (15.6-21.6) | 700 | 107 | 15.3 (12.6-18.0) |
| Regularly using Juul when not allowed to smoke[2] | 636 | 9 | 1.4 (0.5-2.3) | 700 | 5 | 0.7 (0.1-1.3) |
| Using any of the following – e-cigarette[3] | 3667 | 110 | 3.0 (2.4-3.6) | 4131 | 209 | 5.1 (4.4-5.8) |
| Using any of the following – Juul[3] | 3667 | 7 | 0.2 (0.1-0.3) | 4131 | 2 | 0.0 (0.0-0.1) |

[1] Asked to people who smoked in the past year

[2] Asked to people who currently smoke

[3] Asked to people who have not smoke in the past year
